# Neurobiological and clinical effects of High-Definition tDCS on persistent auditory hallucinations in schizophrenia: A randomized controlled trial

**DOI:** 10.1101/2023.05.10.23289796

**Authors:** Rujuta Parlikar, Harleen Chhabra, Sowmya Selvaraj, Venkataram Shivakumar, Vanteemar S. Sreeraj, Damodharan Dinakaran, Satish Suhas, Janardhanan C. Narayanaswamy, Naren P Rao, Ganesan Venkatasubramanian

## Abstract

**Background:** High-definition transcranial direct current stimulation (HD-tDCS) is a potential add-on treatment for persistent auditory hallucinations (AH). However, the lack of evidence from methodical studies implores the need for a systematic evaluation to ascertain its effectiveness.

**Aim:** To examine the clinical and neurobiological role of HD-tDCS in the alleviation of persistent AH and the persistence of its effects in patients with schizophrenia in a double-blinded, sham-controlled study with concurrent resting state fMRI data.

**Methods:** Thirty-four patients with persistent AH were randomized into a TRUE or SHAM arm for five days of the RCT phase (with concurrent resting state fMRI imaging data at baseline and post-RCT), followed by an open-label extension phase of 5 days of TRUE HD-tDCS. In the RCT phase, patients received -2mA current in the TRUE arm and feeble current mimicking sensory effects in the SHAM arm using the 4 × 1 montage at the left temporo-parietal junction (l-TPJ) using subject-specific neuro-navigation. AH severity was assessed using the PSYRATS *Auditory Hallucination Rating Scale* (AHS) at baseline, after RCT, after the end of the open-label, and then by 1^st^ and 3^rd^-month following the last HD-tDCS session. The electric field (EF) was estimated at the region of interest using a simulation technique to further explore the neurobiological effects between the TRUE versus the SHAM group,

**Results:** A significant difference in the neuro-modulatory effect was seen in the neuroimaging analysis at the l-TPJ secondary to the TRUE compared to SHAM HD-tDCS after five days of RCT. At the follow-up, subjects in the SHAM who crossed over to TRUE HD-tDCS significantly improved in AH scores compared to patients who received ten days of TRUE HD-tDCS (T=2.95, p<0.05). However, there was no significant difference in AH scores between the TRUE and SHAM arm at the end of 5 days of RCT and immediately after five days of additional open-label HD-tDCS. In the simulation analysis, differences were noted in the TRUE (EF= 0.22 V/m) versus the feeble current SHAM arm group. It was interesting to observe, that though feeble in magnitude, SHAM current also created a local electric field (EF= 0.007 V/m).

**Conclusions:** Five days of TRUE cathodal HD-tDCS administered to alleviate AH causes cortical effects of interest. Neuromodulatory effects preceded by clinical effects suggest possible clinical latency. Significant improvement in SHAM succeeding TRUE HD-tDCS compared to the ten days of TRUE HD-tDCS suggests the possibility of long-term effects of HD-tDCS acting through mechanisms like homeostatic meta-plasticity. Additional evidence in support of the probable priming effects is the ROI-based electric field simulation showing the generation of local electric field secondary to feeble current in the SHAM arm. Hence sham current with low EF when followed by TRUE current with higher magnitude EF showed enhanced inhibition as compared to the group that followed 10 days of TRUE current further supporting homeostatic meta-plasticity mechanisms.

## Introduction

### Background

Schizophrenia as a disorder has a prevalence of 0.28% of the population (Charlson et al., 2018). One of the target symptoms of this disorder, which is present in a majority of patients, is auditory verbal hallucinations (AH) in about 74% of the patients with schizophrenia (Mueser et al., 1990; Sartorius et al., 1986; Silbersweig and Stern, 1996; Wible et al., 2009). AH is instrumental in substantially relegating the sufferer’s quality of life (Hayward et al., 2017). Interestingly, it has been found that, despite well-monitored antipsychotic treatment, many patients fail to respond adequately, with about 20-30% of patients not obtaining any reduction in their psychotic symptoms (Nieuwdorp et al., 2015; Slotema et al., 2013).

While examining the observations at the cellular level in patients with schizophrenia, there is evidence for dysfunction in the neural plasticity mechanisms (Bhandari et al., 2016). Studies have noted aberrant glutamatergic and GABAergic neurotransmission (Voineskos et al., 2013) which are said to play a role in the long-term potentiation and long-term depression like neuroplastic mechanisms (Hebb, 2005) in these patients. Particularly patients with AH have been found to be associated with higher levels of glutamate and glutamine as compared with non-AVH patients diagnosed with schizophrenia (Weber et al., 2021).

Under clinical realms, parallel and novel treatment options like transcranial direct current stimulation (tDCS) (Slotema, Aleman, Daskalakis, & Sommer, 2012) have been found to alleviate auditory verbal hallucinations significantly(Bose et al., 2019)(Wible, Preus, & Hashimoto, 2009). HD-tDCS is a technique that functions on the same principle as conventional tDCS but is presumed to be more focal (Datta et al., 2009). There is abundant literature investigating the role of conventional tDCS as an intervention in these patients; however, there is limited data on the potential effect of HD-tDCS on auditory hallucinations in schizophrenia. Also, most studies have looked at the immediate effects, not the sustained effects of this intervention.

At the cellular level, the neuroplasticity inducing effects of tDCS have been examined across studies (Hasan et al., 2013). Glutamate NMDA receptors which are known to regulate the firing of the GABAergic neurons, have been noted targets of tDCS (Ghanavati et al., 2022). As HD-tDCS in-principle similar to tDCS (Reckow et al., 2018), however with more focal delivery, it’s interesting to understand its probable role in neuroplasticity and the subsequent clinical translation to alleviation of AVH in schizophrenia.

Interestingly there have been studies that have attempted to understand if inherent plasticity mechanisms can be exploited through NIBS techniques for better regulation of neurotransmission in conditions where it is aberrant (Hurley and Machado, 2017). One of the meta-plasticity phenomena called the homeostatic meta-plasticity has been widely examined in earlier studies (Besson et al., 2016; Opie and Cirillo, 2017; Ziemann and Siebner, 2008). Whether succeeding current intensity enhances or reduces local electric field magnitude based on the strength or polarity of the preceding neuromodulation, and whether this mechanism can be tapped to cause enhanced intended effects at the target site is something that has gathered interest (Opie and Cirillo, 2017).

Examining clinical changes following HD-tDCS helps in understanding its clinical efficacy, whereas evaluating the neural correlates throws light on the mechanism of action. This information may be pivotal in designing appropriate study designs, montage configurations, current intensity parameters, and session durations. Functional neuroimaging analysis might enable us to draw inferences regarding the cortical and subcortical effects of this technique.

In addition, evaluating the long-term clinical effects of HD-tDCS using longitudinal study designs is clinically more relevant than acute effects reported in cross-sectional designs. This will help gather more data on the lasting effects of HD-tDCS and thereby the methods of augmenting its effects. There is a need to investigate the duration for which the improvement (if any) persists in a patient. Most of these studies have not looked at the long-term benefits of this treatment. Indeed, incorporating a follow-up component in controlled trials might help gather more data on the lasting effects of this technique and, thereby, the methods of augmenting its effectiveness.

Therefore, in this study, an attempt has been made to address these compelling arguments. This study uses a neuronavigation-based approach, thereby assuring the specificity of HD-tDCS administration for understanding region-based neuromodulation effects in a randomized, double-blinded fashion for both clinical and neural effects. Since imaging data is available before and after HD-tDCS administration, this study comments on this technique’s effects on neural correlates. This study aims at systematically exploring the sustained clinical and neural effects of HD-tDCS in AVH using a randomized double blind sham-controlled study design with a follow-up component. Further an attempt has been made to understand the mechanistic basis of HD-tDCS using functional neuroimaging and exploring the electric field through neuro-simulation in TRUE compared to SHAM groups. The study objectives were to examine the neural correlates of persistent auditory verbal Hallucinations in schizophrenia patients in comparison with healthy controls and to study the efficacy of add-on HD-tDCS as a treatment modality in these patients. The primary hypothesis was that the neural correlates of persistent AVH will involve frontotemporal, parietal brain regions (as assessed using functional and structural MRI) and that in patients with add-on “true” HD-tDCS, there will be significant improvement immediately in the magnitude of reduction in the severity of auditory verbal hallucination. Such changes will not be seen in schizophrenia patients that receive add-on “sham” HD-tDCS. The secondary hypothesis was that significant differences will be observed in the adaptive modulation (activation and or attenuation of the neural connectivity/activation) pre and post HD-tDCS procedure in subjects with “true” add-on HD-tDCS as compared to subjects who receive add-on “sham” HD-tDCS.

## Materials and Methods

### Patient description

After obtaining ethical approval from the institutional committee, patients attending the clinical services of the National Institute of Mental Health & Neurosciences (India) who fulfilled DSM-5 (American Psychiatry Association (2013)) criteria for schizophrenia (N=34; Age=30.79±7.64 years; M:F=19:15) were diagnosed independently by two psychiatrists. Patients with persistent auditory hallucinations [defined by The Psychotic Symptom Rating Scale (PSYRATS) auditory hallucination sub-scale (Haddock, McCarron, Tarrier, & Faragher, 1999) items of frequency, duration, and disruption each having a score ≥2 despite treatment with adequate antipsychotic dosage for at least 3-months], were recruited in the study. Patients who were left-handed or had any psychiatric emergency, or were pregnant or had any contraindication to procedures like MRI were excluded from the study.

### Study design

Patients were subjected to add-on HD-tDCS treatment using a randomized, double-blind, parallel-arm HD-tDCS protocol.

### HD-tDCS procedure

HD-tDCS procedure was done using standard equipment (https://soterixmedical.com/). Adequate information regarding the procedure was provided to the patients (Villamar et al., 2013). TRUE and SHAM HD-tDCS procedures were performed as per established guidelines with stringent safety standards. The current strength for TRUE HD-tDCS stimulation was (−)2 mA at the cathode and (+)0.5 mA at each anode for 20 minutes. Sham HD-tDCS stimulation level was +0.02mA at the central electrode and three return electrodes, while one electrode was automatically set to return current of -0.08 mA. The sessions were conducted twice a day for five consecutive days with an intersession interval of 3 hours (Table 4). The placement of electrodes was cathode placed at CP5, targeting the left TPJ area [BA 40], and the four returning electrodes (anodes) were placed at FC3, FT7, P1, and P7 as per EEG 10/10 system (Koessler et al., 2009). The precise location of the cathode for all the patients was obtained through a subject-specific MRI-guided neuronavigation approach (https://www.localite.de/en/home/).

### Clinical assessment and outcome

The diagnosis of schizophrenia was established using a structured clinical interview. The capacity to consent was examined using The University of California, San Diego Brief Assessment of Capacity to Consent-UBACC (Jeste et al., 2007). Primary outcome, the severity of auditory hallucinations was measured using the auditory hallucinations subscale (AHS) of PSYRATS (Haddock et al., 1999) on day 1 and day 5 of HD-tDCS. The secondary outcome, the severity of positive and negative symptoms was assessed using Scale for Assessment of Positive Symptoms (SAPS) (Andreasen, 1984) and the Scale for Assessment of Negative Symptoms (SANS) (Andreasen, 1989) at baseline, after 5 days RCT phase. For exploratory analysis, AH, SAPS and SANS were repeated at the end of additional 5 days of open label trial, and additionally the change in AH was examined up to the end of 1^st^ and 3^rd^ month follow up also.

### Statistical analyses

#### Sample size estimation

For evaluating the above-mentioned primary objectives, the optimal sample size was estimated using the principles & methods used by Faul et al. (Faul et al., 2007). Based on new data on the effect of HD-tDCS on auditory verbal hallucinations in schizophrenia (Sreeraj et al., 2018), the power analysis was repeated. As per this re-analysis, the effect size was found to be 1.02; and a sample size of 13 patients in each group was found to be sufficient to detect clinical effects (90% power). This sample was also adequate to detect neurobiologically significant effects on fMRI data as per the recommendations (Desmond and Glover, 2002).

In this design (see Figure 1), the patients as well as the experimenter/rater, were blinded to the randomization of treatment assignment (‘TRUE’ versus ‘SHAM’ add-on HD-tDCS) throughout the study (including a 3-month follow-up). A researcher (uninvolved in the study) apriori created study codes by randomization using a computerized randomization algorithm/ software ascertaining TRUE and SHAM arm subject codes into 1:1 allocation ratio. The codes were opened only after the recruitment of the subject. After phase 1 of RCT (5 days), patients were offered another course of add-on “true” HD-tDCS for five days (while documenting the percentage change of AH scores from baseline, with >20% change in AH scores considered as clinical improvement (Leucht et al., 2006)). The long-term effect of the cortical stimulation by HD-tDCS on the persistent AH was evaluated by conducting a clinical assessment follow-up at 1 and 3 months from the last HD-tDCS session of the patient (see Figure 1).

**Figure 1.**
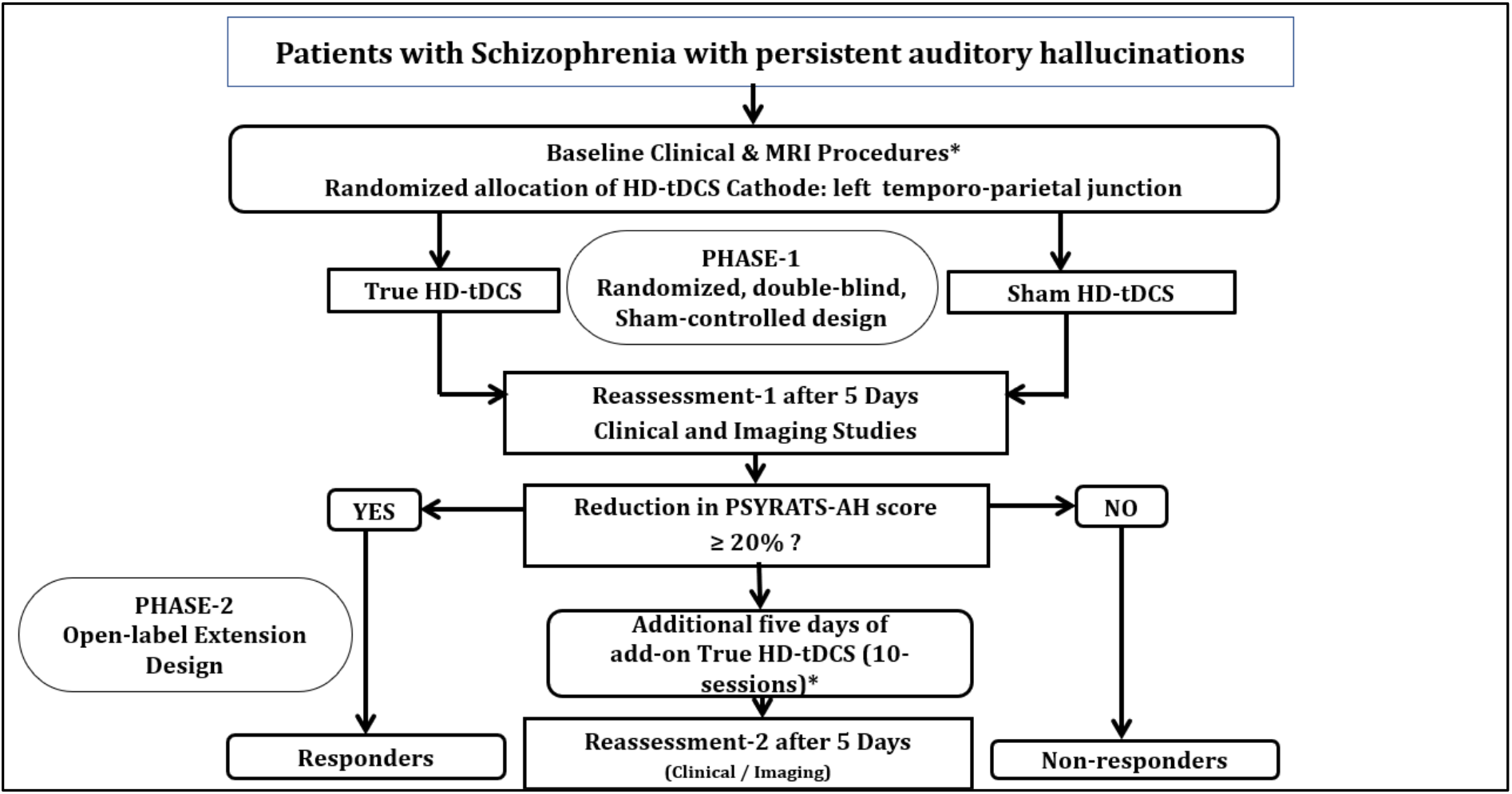
Pictorial representation of the RCT HD-tDCS study design

#### Clinical data

Statistical analysis was done using Linear mixed models analysis of covariance, with age, sex, and olanzapine equivalents as covariates, to examine the effect of add-on HD-tDCS on AH scores and SAPS and SANS score by analysing the interaction effects between the fixed effects of the type of stimulation (TRUE and SHAM) and random effects (i.e., the subjects) at baseline and post RCT HD-tDCS intervention, and effects between the fixed effects of the type of stimulation (TRUE + TRUE and SHAM + TRUE) and random effects (i.e., the subjects) at baseline, post-RCT HD-tDCS and post-open-label TRUE HD-tDCS intervention for non-responders after the RCT phase. Additionally, linear mixed models ANCOVA was also created to look for the interaction effects between the fixed effects of the type of stimulation (TRUE + TRUE and SHAM + TRUE) and random effects (i.e., the subjects) for all subjects who received open label trial after RCT at baseline, post-RCT HD-tDCS, post-open-label TRUE HD-tDCS intervention, at 1-month F/U and 3^rd^-month F/U.

#### Neuroimaging data

MRI data was acquired using a 3T scanner (Ingenia CX, Philips Healthcare, Best, Netherlands, R 5.3.1.2) TR = 6.5 msec; TE = 2.9 msec. The resting-state fMRI data was acquired using BOLD [Blood Oxygen Level Dependent] sensitive echo-planar imaging, yielding 275 dynamic scans. The scan parameters were TR = 2200ms; TE = 28 ms; All the subjects were asked to keep their eyes open during the resting-state fMRI acquisition.

The data were pre-processed and analysed using SPM12 in CONN 20b (Whitfield-Gabrieli and Nieto-Castanon, 2012) on MATLAB R2018b (2018) Apriori Region of Interest (ROI) based analysis was performed based on previous studies looking at auditory hallucinations in schizophrenia (Zmigrod et al., 2016). Significant seed-to-voxel connectivity clusters were identified with a voxel threshold of 0.001 and cluster threshold of 0.05 (FDR corrected).

TPJ was identified as the region between labelled ROIs (Harvard-Oxford (https://fsl.fmrib.ox.ac.uk/fsl/fslwiki/Atlases) atlas-based in CONN) between the four return electrodes as the electric field is distributed within this circumference based on return electrode positions of the 4 × 1 HD tDCS montage. These were the superior temporal gyrus and Heschl’s gyrus (FT7), middle frontal gyrus (FC3), lateral occipital gyrus (PO7), Precuneus (P1), precentral gyrus (FC5), postcentral gyrus and superior parietal lobule (CP1), inferior parietal lobule (P3), inferior temporal gyrus (P7), middle temporal gyrus (TP7) and (T7) surrounding the location of the central electrode (Datta et al., 2009; Koessler et al., 2009),

The effects of HD-tDCS on cortical changes were examined by performing resting-state fMRI connectivity analysis using Repeated measure ANCOVA (RM-ANCOVA) (age, gender, olanzapine equivalents as covariates) with HD-tDCS stimulation type (TRUE vs. SHAM). The correlation was also done to find the associations between changes in clinical scores and hemodynamic changes for the significant connectivity clusters.

### Electric field simulation

Using Automated Neuroimaging Tools (ANTs) software (http://stnava.github.io/ANTs/), a subject-specific template was separately created for the patients in the TRUE group and patients in the SHAM group. After the creation of the template, SIMNIBS software (https://simnibs.github.io/simnibs/build/html/index.html) was used to generate electric field intensity separately for the template from the TRUE group and SHAM group.

In the TRUE group, the parameters for simulation were set as per the stimulation values as follows:

The table demonstrates that there was a net current of 2 mA, with cathodal CP5 (−2.0 mA), and cumulative anodal current of 2 mA-FT7 (+ 0.5 mA), FC3 (+ 0.5 mA), P1 (+0.5 mA), PO7 (+0.5 mA)

In the Sham group (see supplementary Figure 1), four electrodes were set (machine default) to receive 0.02 mA of current with the 5^th^ electrode carrying the return current of -0.08 mA. To ascertain that l-TPJ received the lowest current during SHAM treatment, the central electrode at CP5 was set to receive +0.02 mA of constant current in all the patients. However, the position of the electrode carrying the return current of -0.08mA was randomly randomized across the patients in the SHAM group. Hence patients in the SHAM group received (with a random probability of receiving any) one of the any four possible montages for 10 sessions.

It is imperative to mention that patients in the sham group could have received different sham configuration during different sessions of the RCT arm since the position of the return electrode (−0.08 mA) was randomly randomized (See Figure 1 supplementary).

## Results

### Prospective component (RCT-HD-tDCS)

#### Recruitment

Patients were recruited from 2018 March to 2019 October. All patients were followed up post 1 month and post 3^rd^ month of the last session of HD-tDCS session (see Figure 2).

**Figure 2.**
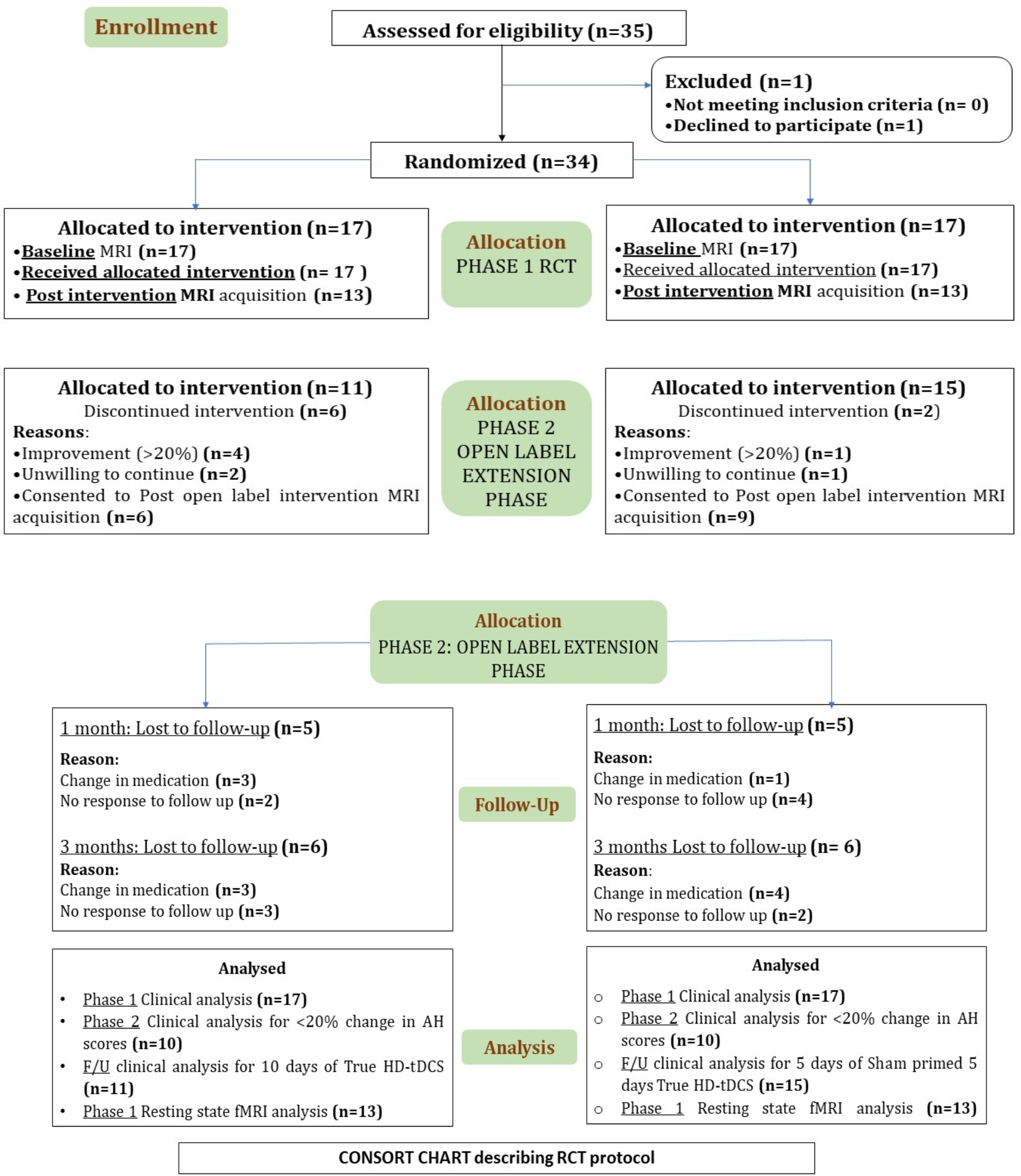
CONSORT flowchart describing the enrolment, data acquisition and analysis

#### A comparative profile of patient characteristics

The study sample had 34 patients (N=34; Age=30.79±7.64 years; M: F=19:15), with 17 in the TRUE arm (N=17; Age=31.71 ± 7.48 years; M: F=10:7; Olanzapine equivalents=45.01 ± 76.90) and 17 in the SHAM arm (N=17; Age=29.88 ± 7.92 years; M: F=9:8; Olanzapine equivalents=57.33 ± 86.33).

As age (Cohen, Izediuno, Yadack, Ghosh, & Garrett, 2014), gender (Suessenbacher-Kessler et al., 2021), and antipsychotics (Johnsen, Hugdahl, Fusar-Poli, Kroken, & Kompus, 2013) are likely to contribute to the baseline as well as change in the symptom scores with HD-tDCS, spearman correlation was done to establish any associations among them. No significant correlation was observed among the variables.

#### Observations on PSYRATS-AHS during

##### RCT [5 days of TRUE or 5 days of SHAM HD-tDCS]

Using a linear mixed model approach, after controlling for age (mean 30.79 ± 7.64), sex (M: F=19:15), and olanzapine equivalents (mean 51.17 ± 80.74) for all the patients (n=34), the HD-tDCS group × time interaction was not found to be significant (T= 0.81 and p=0.42) for AH scores (95% CI [-3.29, 7.70]) (See Supplementary Section Figure 2)

#### Effect of HD-tDCS on resting-state fMRI measures of AH in SCZ

Out of 34 patients, neuroimaging data (baseline and post RCT) was available for 26 patients at the end of phase 1 (since seven patients declined to re-undergo MRI and one patient post-RCT fMRI data was of suboptimal quality secondary to significant motion artifacts). With repeated measures ANCOVA model, significant differences in the connectivity after controlling for age (mean 30.23± 7.95), sex (M: F= 14:12), and olanzapine equivalents (mean 32.95 ±38.13) were observed for all the patients in TRUE (n=13) and SHAM arm (n=13)

There was decreased connectivity (see Supplementary section Figure 3 and Table 1) between the left Heschl’s gyrus and the left and right caudate (see Supplementary section Figure 3a) after TRUE HD-tDCS compared to Sham HD-tDCS. In the TRUE HD-tDCS arm, there was decreased connectivity between the left lateral occipital cortex superior division and the Precuneus cortex (see Supplementary section Figure 3b). Increased connectivity was observed between the left lateral occipital cortex superior division with right lateral occipital cortex inferior and superior division (see Supplementary section Figure 3c) and between the left lateral occipital cortex inferior division and the left lateral occipital cortex superior division and superior parietal lobule (see Supplementary section Figure 3d).

**Table 1:** Stimulation parameters in TRUE group

|  |  |  |
| --- | --- | --- |
| Central electrode | (CP5)- corresponding to the I-TPJ | - 2mA |
| Return electrodes | FT7, FC3, P1, PO7 | + 0.5 mA |

#### Correlation between the change in connectivity clusters (Δ of beta values) after HD-tDCS with percentage change in AH (mean 18.55 ± 20.61) scores

Among the three clusters, left iLOC connectivity with left sLOC and left SPL showed a significant negative correlation within the SHAM group (n=13) patients (rho= -0.88, p= 0.002) (see Supplementary section figure 4). No correlation was observed within the TRUE group patients.

##### Secondary outcome: RCT [5 days of TRUE or 5 days of SHAM HD-tDCS]

The HD-tDCS group × time interaction was not found to be significant for SAPS (95% CI [-6.72, 10.94]) and SANS scores (95% CI [-15.54, 4.65]).

##### Open-label

Non-responders from RCT (<20% improvement in AH scores from baseline)

###### Primary outcome

Sub group analysis was done to examine if the open label trial with HD-tDCS had any effect on patients who did not respond to RCT phase. 23 patients out of 34 showed <20% improvement in AH scores from baseline at the end of phase 1 RCT. 20 patients out of those 23 patients received another 5 days of TRUE HD-tDCS treatment. Using a linear mixed model approach, after controlling for age (mean 31.45 ± 8.24), sex (M: F, 12: 8), and olanzapine equivalents (mean 52.89 ± 78.66), there was a significant decrease in the AH scores from baseline to post HD-tDCS in TRUE + TRUE arm (baseline C.I. 31.70 to 35.26, and post-C.I. 22.80 to 29.58) as well as in SHAM arm (baseline C.I. 29.64 to 33.34, and post-C.I. 21.93 to 28.84). However, the HD-tDCS group (TRUE + TRUE or SHAM + TRUE) × time interaction was not significant (T=0.3, p=0.7) for AH scores.

###### Secondary outcome

The HD-tDCS group × time interaction was not found to be significant for SAPS (95% CI [-8.46, 10.5]) and SANS scores (95% CI [-15, 6.59]).

###### All patients who received open label trial (n=26)

Follow up analysis was done to examine the effects of add-on TRUE HD-tDCS (TRUE + TRUE or SHAM + TRUE) in all patients (n=26) who received additional open label trial irrespective of their improvement status after the RCT phase at one month and at the 3^rd^-month. 26 patients out of 34 received the open-label trial. In linear mixed-models analysis the HD-tDCS group (TRUE + TRUE or SHAM + TRUE) × time (baseline, RCT and post-open-label, one monthly F/U and 3 monthly F/U) interaction for AH symptom scores after controlling for age (mean 31.04 ± 8.12), sex (M: F, 16:10) and olanzapine equivalents (mean 49.38 ± 69.51) was done.

After controlling for the main effects of age, sex, and olanzapine equivalents, the group-by-time interaction was statistically significant for AH scores (T=2.95, p<0.001). There was a significant decrease in the AH scores (see Figure 3) from baseline to post-intervention with HD-tDCS in the SHAM +TRUE arm (*baseline C*.*I. 30*.*24 to 33*.*30, and 3rd monthly C*.*I. 7*.*89 to 20*.*62*) whereas in TRUE + TRUE was not found to be significant (*baseline C*.*I. 31*.*86 to 34*.*93, and 3^rd^ monthly C*.*I. 20*.*49 to 34*.*52*) at 3^rd^-month follow-up.

**Figure 3.**
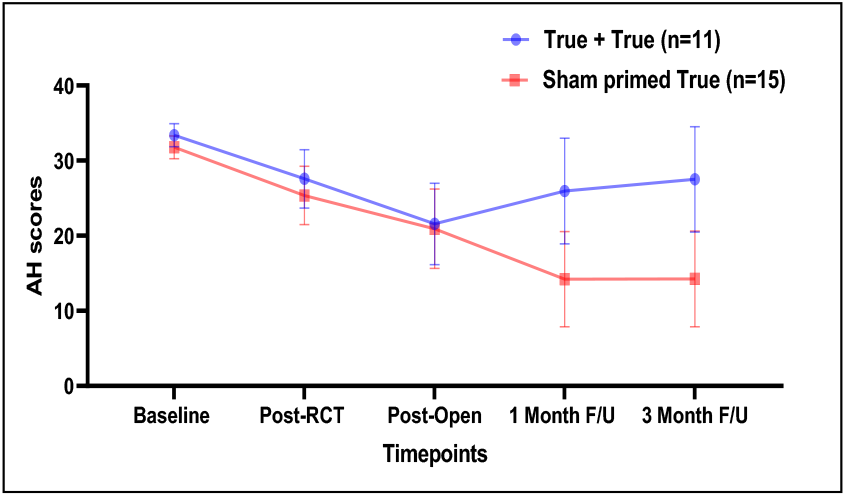
There was a significant reduction in the mean AH scores for sham + TRUE HD-tDCS group compared to 10 days of TRUE HD-tDCS group at F/U 1 month and 3^rd^ month after the last time point of TRUE HD-tDCS stimulation in phase 2 of the post-open label.

**Figure 4.**
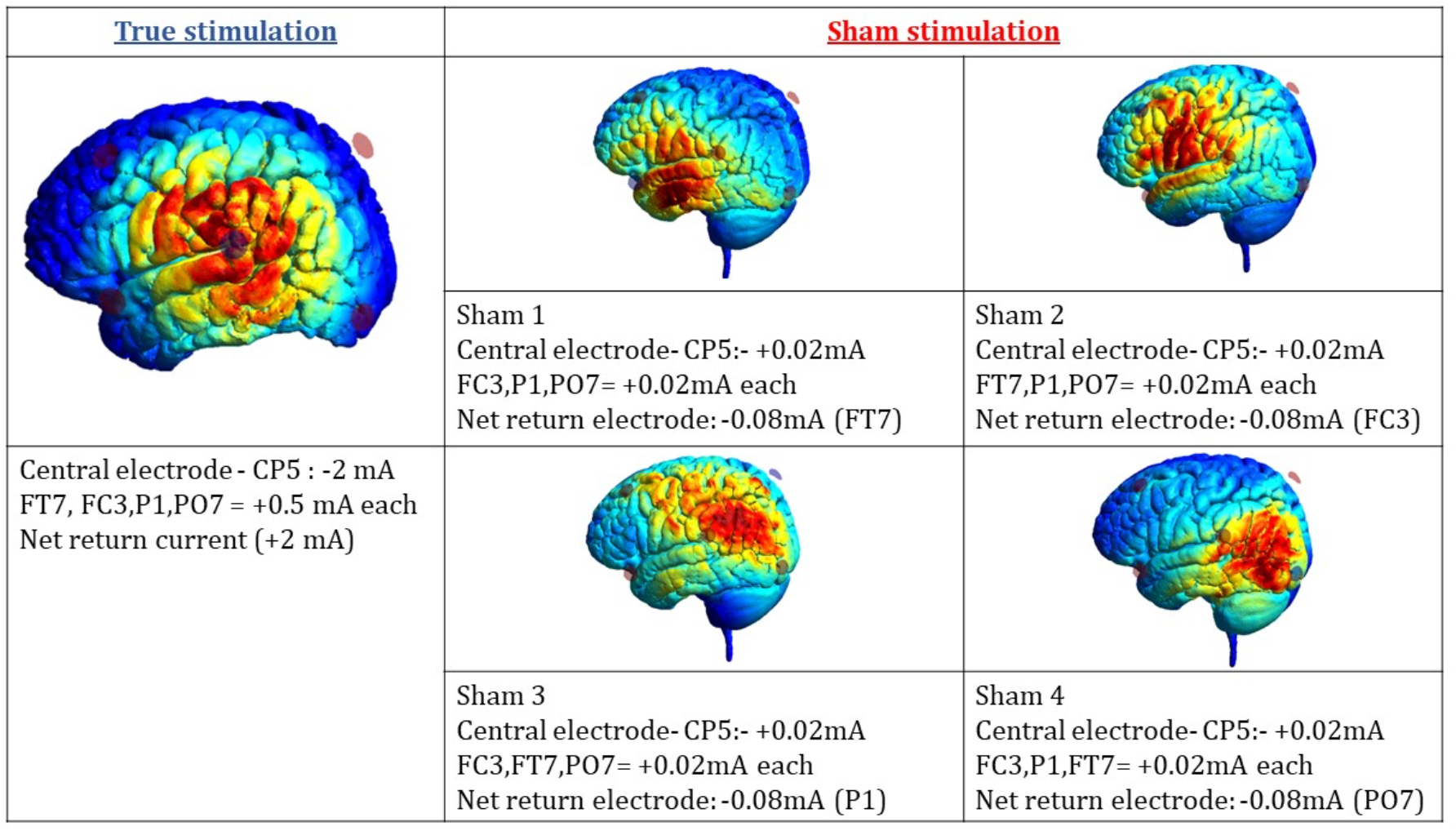
The pictorial representation of the EF for both the groups is described. In order to determine the local EF intensity, ROI based analysis was done using the SIMNIBS matlab script. When the local average EF was examined under CP5 (corresponding to l TPJ), the following results were noted

#### An error plot of mean and SE for AH scores in TRUE + TRUE and SHAM primed TRUE

##### Electric field distribution

For TRUE group versus SHAM group

Five different simulations were run (see Figure 4); one simulation on the subject specific template for the true group and four different simulations were run on the subject specific template for the Sham group. The results are illustrated below:

It was observed (See Table 2-Supplementary material) if the patient belonged to the TRUE group, there was 0.25 V/m EF per session, while if he/she belonged to the SHAM group, there was on an average 0.007 V/m per session at the l-TPJ ROI region. This analysis also suggests that patients who received 10 days of TRUE received a dose of 0.25 V/m EF every session for 10 days while those who switched from SHAM to TRUE received 5 days of low EF of 0.007 V/m per session followed by 0.25 V/m of EF per session at their l-TPJ during the open-label True phase for the next 5 days.

**Table 2:**
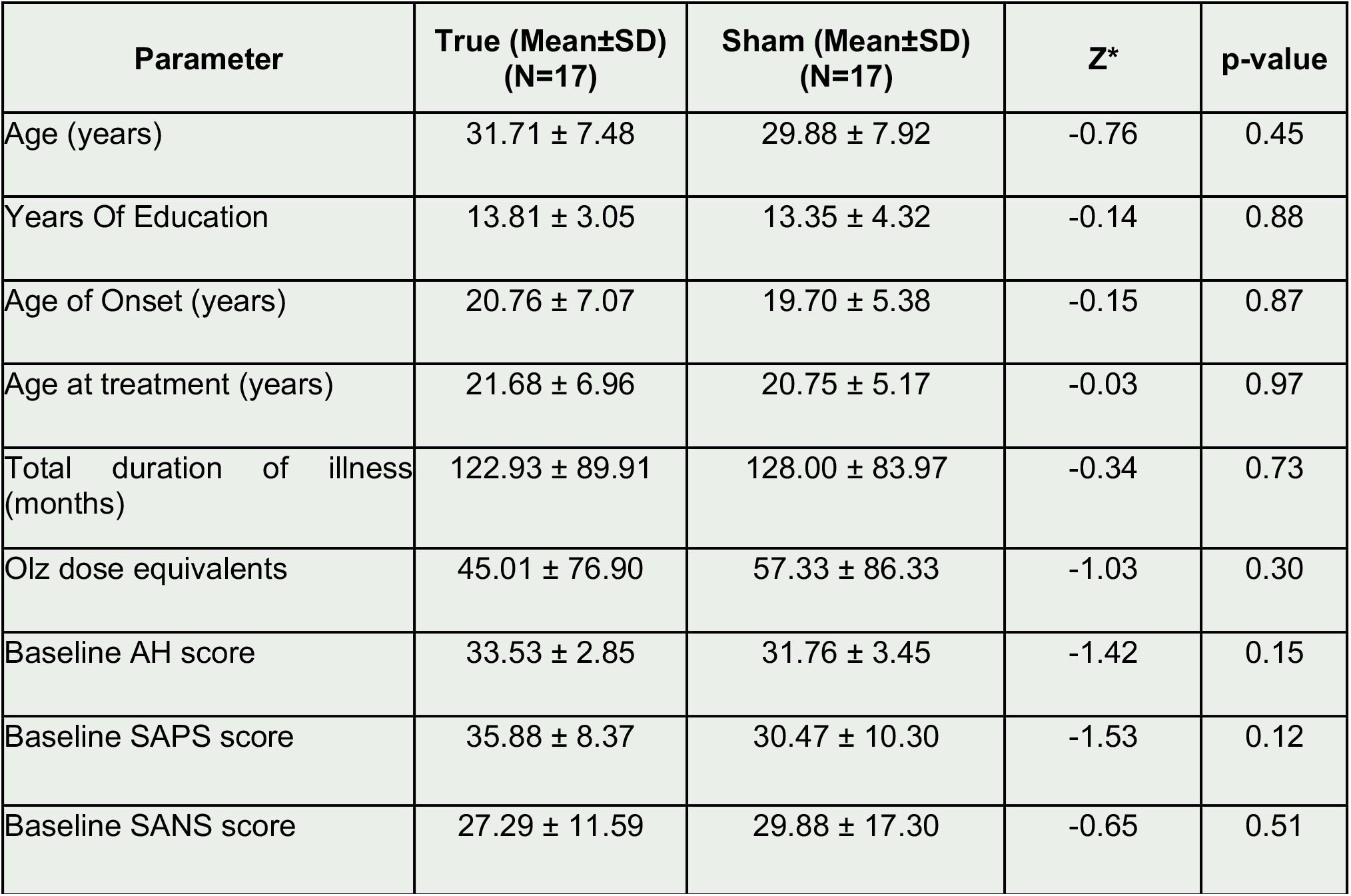
Baseline demographic and clinical characteristics of each group. All the parameters are equally distributed among the two groups (True and sham group) *compared using Mann Whitney U-test.

These results also suggest that irrespective of which combination of sham montage the patient in the sham group during phase 1 of RCT would have received, there was an average of 0.007 V/m EF at the left TPJ area.

## Discussion

There was a significant reduction in the magnitude of the AH scores after intervention with five days of add-on HD-tDCS. Although group-by-time interaction was not significant, there was a significant change in the objective measures of resting-state connectivity between the two groups. One of the possible reasons for this discrepant observation can be understood based on the findings in the resting change fMRI measures after HD-tDCS.

In the Apriori seed-to-voxel connectivity analysis, it was observed that there was a significant reduction in the connectivity of the Heschl’s gyrus (l-HG) with the left and the right caudate in the TRUE group versus the SHAM group. HG has been consistently implicated in the pathophysiology of AH (Dierks et al., 1999). The caudate nucleus, an integral part of the basal ganglia, has also been suggested to play a role in the pathogenesis of AH (Scheepers, Gispen de Wied, Hulshoff Pol, & Kahn, 2001). A significant reduction in the connectivity of this regional communication perhaps suggests that cathodal inhibition at l-TPJ decreases the aberrant hyperconnectivity of the primary auditory cortex with regions that may be pivotal in the generation or modification of hallucinatory experiences.

Another region within the left TPJ, the left superior lateral occipital cortex (sLOC), was found to have decreased connectivity (Zhuo et al., 2016) with the Precuneus (implicated in the AH experience) following TRUE HD-tDCS compared to SHAM HD-tDCS. After TRUE HD-tDCS, there was increased connectivity of left sLOC with contralateral sLOC and increased connectivity of left iLOC with the left sLOC and left SPL. LOC and SPL, considered part of the network critical for multisensory integration, are said to play a role in integrating sensory components in a perceptual experience (Beauchamp, 2005; Giovannelli et al., 2016). Increased connectivity with these regions perhaps suggests a better multisensory integration post-TRUE HD-tDCS. The findings related to occipital cortex align with the increasing evidence for the role of occipital cortex (Xue et al., 2022; Shaw et al., 2020) and visual system abnormalities in schizophrenia pathogenesis (Silverstein et al., 2021) that fit with the overarching postulate of predictive processing deficits (Adams et al., 2013; Sterzer et al 2018) involving imbalanced neuroplasticity (Guterman et al 2021)

Given that, in this study, both TRUE and SHAM stimulation were seen to cause neuro-modulatory cortical effects, there is a substantial possibility that the SHAM stimulation (constant current of +0.02mA at l-TPJ) in this study did not act like a placebo arm but rather a weak active arm capable of inducing cortical effects. And interestingly, only within the SHAM group, the connectivity changes of l-iLOC with l-sLOC and l-SPL were found to be negatively associated with the percentage change in AH, suggesting that patients were less likely to improve with cortical changes secondary to SHAM stimulation administered in this study.

Akin to 5 days of Phase 1 RCT, there was no significant difference between AH reduction after additional five days of TRUE HD-tDCS (total of 10 days) in either of the two groups (TRUE +TRUE and SHAM +TRUE). However, when all the patients, who received additional five days of add-on HD-tDCS, irrespective of their improvement status, were examined at the follow-up of 1 and 3rd months, AH reduction was observed for patients who received five days of SHAM HD-tDCS followed by five days of TRUE HD-tDCS as compared to those who received ten days of TRUE HD-tDCS. Indeed, supposing SHAM stimulation was not a placebo arm but rather capable of causing underlying cortical effects (Nikolin, Martin, Loo, & Boonstra, 2018), then the subsequent five days of TRUE cathodal inhibition in the SHAM +TRUE arm may have led to clinical effects on account of mechanistic effects of homeostatic metaplasticity a neural mechanism that inherently adjusts the modulated threshold to preserve the neuron’s activity in the physiologically normal range (Bocci et al., 2014).

To understand the possibility of priming effects, simulation analysis was attempted in this study. It was found that patients who received sham stimulation had a near constant weak EF at the left temporo-parietal junction during all 5 days of stimulation. Hence, when these patients were switched to the TRUE treatment, they inherently received a strong cathodal inhibitory current (with 0.25 V/M at the left TPJ) for another 5 days. Given the clinical results of significant improvement in this group could then be attributable to the possible priming effects at the left TPJ by weak current followed by strong inhibitory cathodal current. In the group that received 10 days of TRUE treatment however, perhaps the collective effects of strong inhibitory stimulation were offset by the homeostatic mechanisms corroborating with the clinical scores at the end of the 1^st^ and 3^rd^ month in the 10 days True treatment group.

Based on the principle of homeostatic metaplasticity, the neuronal threshold at the left TPJ in the TRUE group could have been modulated such that it was less likely to get inhibited if followed with another session of 2 mA cathodal current during an open-label trial. And in the group receiving weak SHAM stimulation, the threshold to get inhibited with the second course of cathodal current would have been modulated less, leading to increased inhibition with subsequent 2mA cathodal current during the open-label trial.

Thus, in this study, the clinical improvement in AH alleviation presumably occurred secondary to cathodal current at left TPJ, as suspected. However, the apparent absence of clinical effects in the TRUE +TRUE group perhaps was secondary to the homeostatic metaplasticity mechanism that counteracted the cumulative effects of the 10 days of cathodal inhibition at the left TPJ (Lang et al., 2004) as against five days in the SHAM + TRUE group. Also, given that the apparent homeostatic effects were not noticeable immediately after the 10th day of HD-tDCS, the changes were more likely to be slow and long-lasting than immediate.

In conclusion, this randomized doubled blinded parallel-arm study demonstrates that 5-days of twice daily, 20-minute sessions of HD-tDCS are sufficient to cause proposed neural effects (as demonstrated by fMRI connectivity changes) but not significant to reflect immediate clinical effects. This study has demonstrated that the weak current primed cathodal inhibition of the left TPJ is beneficial in a significant reduction of AH from baseline compared to cumulative ten days of cathodal stimulation. Also, these effects are seen not immediately but over some time, suggesting latent effects and inherent homeostatic metaplasticity mechanisms of the brain.

To the best of our knowledge, this is the first RCT study to report the clinical effects of HD-tDCS in patients with schizophrenia and changes with the intervention on the resting-state fMRI in a randomized controlled trial. This is the first study looking at the long-term benefits of HD-tDCS stimulation by examining the patients in a follow-up while commenting on priming effects secondary to homeostatic metaplasticity. Additionally, this study used subject-specific neural targeting using neuronavigation protocol to facilitate treatment administration specificity. Limitations of the study include the high expectancy effect with the current sample size and the novelty of the intervention. Also, in this study, the participant’s belief of the TRUE or SHAM stimulation was not examined after the RCT, which could have facilitated in studying the placebo effects, if any.

In future studies, using very low current stimulation in tDCS or HD-tDCS, in comparison with a proper placebo arm and an active TRUE arm, the role of ‘very weak’ versus ‘weak’ versus ‘no current’ in inducing neuro-plasticity can be examined more systematically. Mechanisms acting through homeostatic mechanisms like the effects of weak current primed cathodal intervention can be explored. And lastly, following neuro-modulatory interventions like HD-tDCS, obtaining neuroimaging data at all timepoints of clinical data acquisition can be acquired for corroboration of the clinical findings with the neural correlates to help optimize a better intervention for patients with persistent AH in schizophrenia.

## Other Information

### Registration

CTRI/2018/02/012061-Clinical Trial Registry of India

## Data Availability

All data produced in the present study are available upon reasonable request to the authors

## Acknowledgments

Dr. Rujuta Parlikar is supported by the Department of Biotechnology (DBT)-Wellcome Trust India Alliance (IA/CRC/19/1/610005). Dr. Venkataram Shivakumar is supported by DBT-Wellcome Trust India Alliance Early Career Fellowship Grant (IA/CPHE/18/1/503956). Vanteemar S Sreeraj acknowledges the support of the India-Korea joint program cooperation of science and technology by the National Research Foundation (NRF) Korea (2020K1A3A1A68093469), the Ministry of Science and ICT (MSIT) Korea, and the Department of Biotechnology (India) (DBT/IC-12031(22)-ICD-DBT). Dr. G Venkatasubramanian acknowledges the support of DBT, Wellcome Trust India Alliance (IA/CRC/19/1/610005), and the Department of Biotechnology, Government of India (BT/HRD-NBA-NWB/38/2019-20(6)).

