## Supplementary Tables and Figures for "Neurobiological and clinical effects of High-Definition tDCS on persistent auditory hallucinations in schizophrenia: A randomized controlled trial"

### Supplementary material:

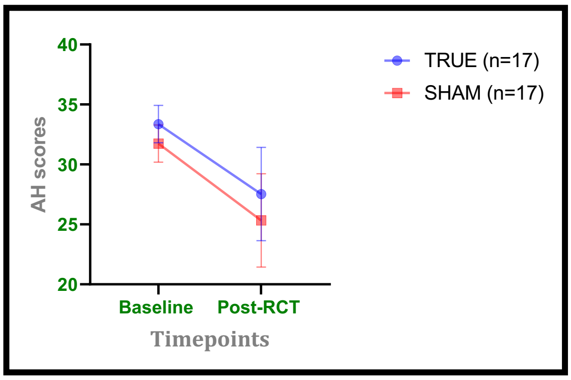


Figure 1: There was a significant reduction in the mean AH scores for both the groups (TR UE and SHAM), but the HD-tDCS group × time interaction was not statistically significant.

Table 1: Tabulation of the significant seed to voxel connectivity clusters for within (post HD>baseline) and between subjects (True>Sham) (n=26)

| - | | - | - | MNI | | | CONN_Localization |
|---|---|---|---|---|---|---|---|
| **Seed** | **P value (FDR corrected)** | **Beta** | **Cluster size** | **x** | **Y** | **z** | **Area** |
| Left Heschl's gyrus | <0.05 | -0.24 | 44 | -6 | 12 | 10 | **Left caudate** (also right caudate) |
| Left Lateral occipital cortex-superior division | <0.05 | -0.26 | 279 | 8 | -64 | 24 | **Precuneus cortex (**also cingulate gyrus posterior division) |
| | <0.05 | 0.33 | 62 | 44 | -82 | 10 | **Right lateral occipital cortex inferior** (also superior right lateral occipital cortex) |
| Left lateral occipital cortex-inferior division | <0.05 | 0.3 | 115 | -34 | -58 | 50 | **Left Lateral occipital cortex superior** (also left superior parietal lobule) |

Table 1: Superior and inferior lateral occipital cortex and Heschl's gyrus show significantly differing connectivity clusters between patients who received TRUE HD-tDCS (n=13) as compared to sham HD-tDCS (n=13) at $P_{FDR} < 0.05$

#### Mean normE in l TPJ ROI:

| **RCT phase – 1** | | | | **Open Label (True Treatment)** | |
|---|---|---|---|---|---|
| | | Average EF per session | | | Average EF per session |
| **TRUE group** | | 0.250 V/m | | **TRUE followed by TRUE** | 0.250 V/m |
| | Stimulation configuration | | Average EF per session | | |
| **SHAM group** | 1 | 0.007 V/m | 0.007 V/m | **Sham group switched to TRUE** | 0.227 V/m |
| | 2 | 0.007 V/m | | | |
| | 3 | 0.007 V/m | | | |
| | 4 | 0.008 V/m | | | |

Table 2 showing the mean average electric field in the left TPJ ROI region after stimulation in TRUE group and SHAM group.

**Neurobiological and clinical effects of High-Definition tDCS on persistent auditory hallucinations in schizophrenia: A randomized controlled trial**

Rujuta Parlikar*, Harleen Chhabra, Sowmya Selvaraj, Venkataram Shivakumar, Vanteemar S. Sreeraj, Damodharan Dinakaran, Satish Suhas, Janardhanan C. Narayanaswamy, Naren P Rao, Ganesan Venkatasubramanian*

Figure 2: Using SIMNIBS software, the simulations were run for both the group specific templates after entering the aforementioned stimulation parameters.

<table>
<tr>
<td>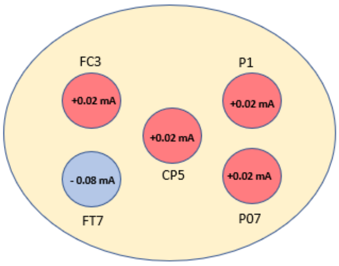
</td>
<td>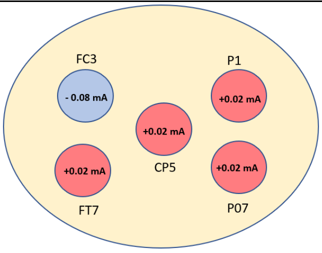
</td>
</tr>
<tr>
<td>CP5 (+0.02 mA)<br>PO7 (+0.02 mA) FC3 (+0.02 mA) P1 (+0.02 mA)<br>FT7 (-0.08 mA)</td>
<td>CP5 (+0.02 mA)<br>FT7 (+0.02 mA) P1 (+0.02 mA) PO7 (+0.02 mA)<br>FC3 (-0.08 mA)</td>
</tr>
<tr>
<td>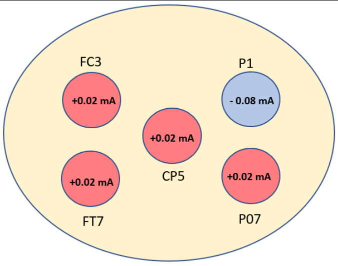
</td>
<td>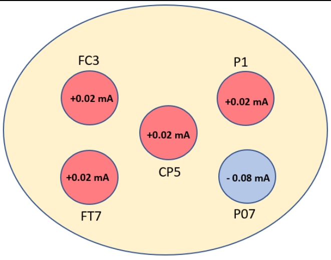
</td>
</tr>
<tr>
<td>CP5 (+0.02 mA)<br>FT7 (+0.02 mA), FC3 (+0.02 mA), PO7 (+0.02 mA)<br>P1 (-0.08 mA)</td>
<td>CP5 (+0.02 mA)<br>FT7 (+0.02 mA), FC3 (+0.02 mA), P1 (+0.02 mA)<br>PO7 (-0.08 mA)</td>
</tr>
</table>

**Neurobiological and clinical effects of High-Definition tDCS on persistent auditory hallucinations in schizophrenia: A randomized controlled trial**

Rujuta Parlikar*, Harleen Chhabra, Sowmya Selvaraj, Venkataram Shivakumar, Vanteemar S. Sreeraj, Damodharan Dinakaran, Satish Suhas, Janardhanan C. Narayanaswamy, Naren P Rao, Ganesan Venkatasubramanian*

Figure 3: An error plot of mean and SE for AH scores in TRUE (n=17) and SHAM arm (n=17) at baseline and
post RCT intervention**.**

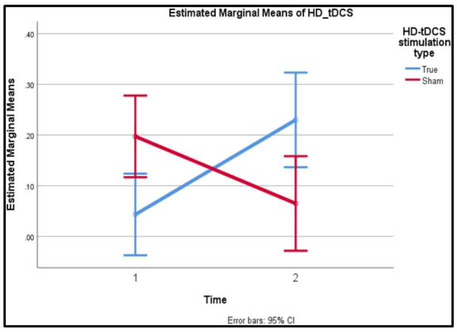


There was an increase in connectivity of left superior lateral occipital cortex (LOC) with right LOC after True HD-tDCS but a reduction in connectivity with sham HD-tDCS.

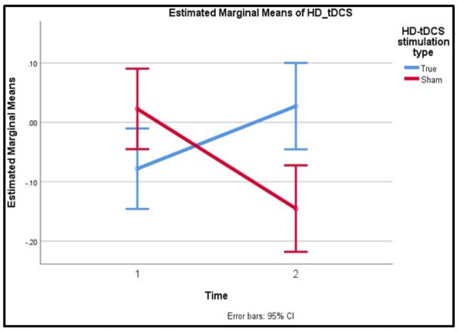


There was an increase in connectivity between left inferior lateral occipital cortex (LOC) with left superior LOC after True HD-tDCS as compared to sham HD-tDCS.

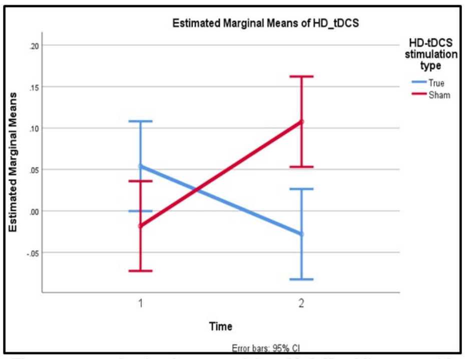


There was a reduction in connectivity of left Heschl's gyrus with the left caudate (and right caudate) after True HD-tDCS but increased connectivity after sham HD-tDCS.

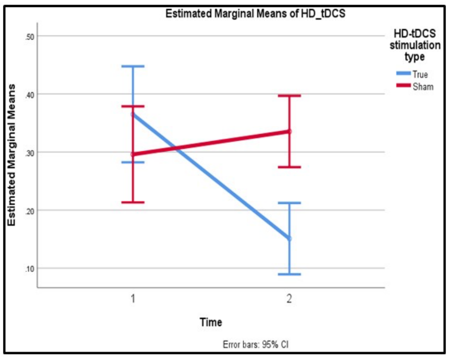


There was a reduction in connectivity of left superior lateral occipital cortex with the Precuneus (and posterior cingulate cortex) after True but not sham HD-tDCS.

Rujuta Parlikar*, Harleen Chhabra, Sowmya Selvaraj, Venkataram Shivakumar, Vanteemar S. Sreeraj, Damodharan Dinakaran, Satish Suhas, Janardhanan C. Narayanaswamy, Naren P Rao, Ganesan Venkatasubramanian*

**Figure 4 There was a significant negative correlation between iLOC connectivity with sLOC_l and SPL_l after sham HD-tDCS, with the change in AH scores (Δ of AH scores after sham HD-tDCS) within the sham group (n=13).**

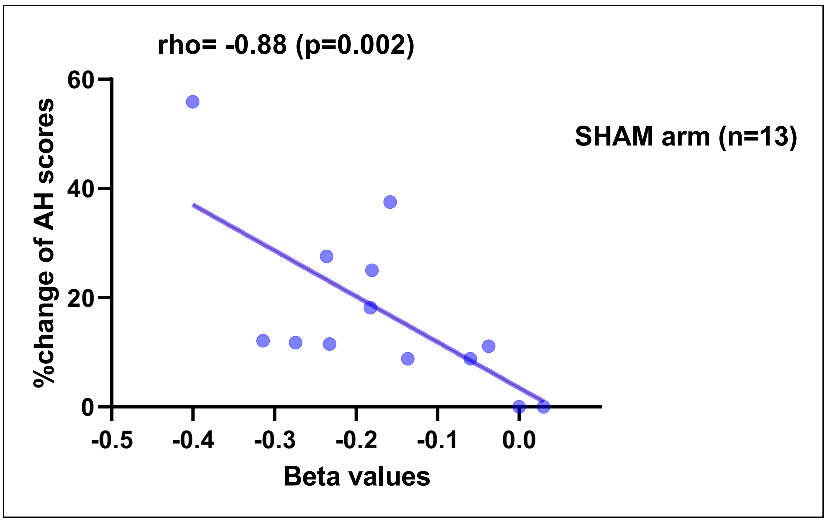
